# PROMIS scales for assessment of the impact of post-COVID syndrome: A Cross Sectional Study

**DOI:** 10.1101/2021.05.25.21257817

**Authors:** Ravindra Ganesh, Aditya K Ghosh, Mark A Nyman, Ivana T Croghan, Stephanie L Grach, Christopher V Anstine, Ryan T Hurt

**Affiliations:** Division of General Internal Medicine, Mayo Clinic, Rochester, MN; Department of Internal Medicine, Mayo Clinic, Rochester, MN; St George’s University, Grenada, West Indies

**Keywords:** PROMIS, post-COVID syndrome, social roles, fatigue

## Abstract

The post-COVID syndrome is estimated to occur in up to 10% of patients who have had COVID-19. This condition manifests as lingering symptoms which persist for weeks to months after resolution of the acute illness. The syndrome is poorly understood and efforts are just beginning to appropriately characterize the symptoms expressed by this population. We present a population of patients with persistent symptoms as measured by a select number of PROMIS surveys (i.e. fatigue, sleep, pain, physical functioning, and social roles). We believe this to be the first use of the PROMIS survey data collected in this population and one of the first to attempt to measure social dysfunction secondary to the post-COVID syndrome. Our patient population is notably younger (30.9% were between 40-59 years of age), with a majority being female (60.5%). They also reported deficits in social roles (34.5%), and greater fatigue (14.7%), and pain (15.9%); along with a variety of disease severity ranging from asymptomatic to requiring admission. Despite this increased heterogeneity of population, the symptomatology of the post-COVID syndrome is preserved. These findings differ significantly from previously published data that demonstrated that outpatients can have duration of post-COVID syndrome similar to those who were hospitalized.

## Introduction

In 2020, the global medical community encountered the generational challenge created by the Severe Acute Respiratory Syndrome Coronavirus 2 (SARS-CoV-2), the causative agent of Coronavirus Disease 2019 (COVID-19) pandemic. This previously unidentified coronavirus infection has been highly contagious, infecting over 150 million people worldwide as of May 2021.^1^ Patients with severe COVID-19 often require hospitalization due to acute respiratory distress secondary to COVID-19 pneumonitis which can proceed to a multisystem inflammatory disease which can necessitate ICU care, mechanical ventilation, and death. As of May 2021, over 3 million deaths have been caused by COVID-19.^1^

Though most patients eventually recover from their acute COVID-19 infection (< 3 weeks), it has now become well recognized that many patients continue to suffer from persistent post-infectious symptoms including fatigue, dyspnea, pain, and palpitations.^2-5^ The prevalence of persistent post-COVID symptoms has been estimated at about 10% in the general population, and may be in excess of 70% in the post-hospital setting.^2,3,6^ As we currently have in excess of 147 million patients who have recovered from acute COVID-19 infection, there are likely several million people suffering from these persistent symptoms. Elucidating the duration, frequency, and type of symptoms after acute COVID-19 infection will provide clarity to the post-COVID syndrome and enable better research into the underlying pathophysiologic mechanisms, thereby guiding individualized patient management.

Given the extreme infectivity of the SARS-CoV-2 virus, public health recommendations have included physical distancing, cancelling of many in person social events, and curtailing of travel. While these measured have slowed the spread of this highly contagious viral illness, there has also been a coincident increase in social isolation, depression, anxiety, and other adverse mental health conditions, including suicidal ideation and substance abuse.^7^ These conditions are likely exacerbated by the financial impact of the pandemic which has left many without jobs and steady income. Assessment of impairments in social roles and adverse impact on mental health could allow for early interventions and improved outcomes.

The Mayo Clinic COVID Frontline Care Team (CFCT) in Rochester, Minnesota has provided telemedicine care for over 35,000 patients who tested positive for COVID-19 in the Midwest since March 2020, including follow up post hospital discharge.^8,9^ This telemedicine team was one of the multiple innovations undertaken by the Mayo Clinic in the COVID-19 response which has led to favorable results.^10^ Many of these patients complained of persistent symptoms post COVID infection, primarily concerning impairments in functional aspects of life. The primary aim of this study was to determine the prevalence and characteristics of persistent physical and social impairment after recovery from acute COVID-19 infection.

## Methods

This retrospective study was approved by the Mayo Clinic Institutional Review Board. The Mayo Clinic maintains a COVID recovered registry which is defined by history of positive PCR for SARS-CoV-2 and meeting the clinical definition of recovery from COVID-19: either resolution of symptoms or >30 days post initial positive PCR test. We identified 7,500 patients from the Midwest region who were on the COVID recovered registry as of September 12, 2020. Of these, 5,604 had an active patient portal in the electronic medical record (EMR) and 4,535 of these had documented research consent. PROMIS (Patient-Reported Outcomes Measurement Information System) questionnaires on fatigue, sleep, social roles, physical function, and pain were sent to these 4,535 patients. A total of 825 (18.2%) patients responded to the surveys. Eight of the respondents did not have complete demographic information in their chart so they were not included in the final analysis which had a total of 817 patients (Figure 1).

**Figure 1:**
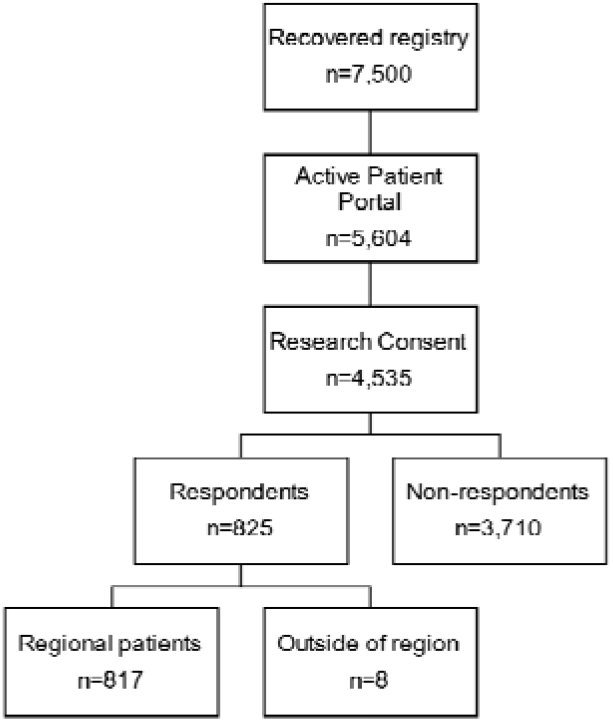
Patient selection and response to questionnaires.

Demographic characteristics were abstracted from the electronic medical record (EMR) and are presented in Table 1. Z-scores and their associated p-values were calculated for these two groups and are also presented in Table 1.

**Table 1:** Demographic characteristics of survey respondents and non-respondents in the recovery cohort.

|  | Non-respondents | Respondents | Z-score | p-value |
| --- | --- | --- | --- | --- |
| Total number of patients | 3710 | 817 |  |  |
| Mean age | 37 (17) | 44 (17) | 11.77 | <0.001 |
| Gender |  |  |  |  |
| Female | 1822 (49.1%) | 499 (61.1%) | -6.19 | <0.001 |
| Male | 1883 (50.9%) | 318 (38.9%) | 6.13 | <0.001 |
| Unknown | 5 (0.1%) | 0 (0%) | 1.05 | 0.29 |
| Primary Language |  |  |  |  |
| English | 3202 (86.3%) | 787 (96.3%) | -8.01 | <0.001 |
| Non-English: Interpreter not needed | 137 (3.7%) | 12 (1.5%) | 3.26 | 0.001 |
| Non-English: Interpreter needed | 295 (8.1%) | 18 (2.2%) | 5.86 | <0.001 |
| Race |  |  |  |  |
| American Indian/Alaskan Native | 6 (0.2%) | 2 (0.2%) | -0.51 | 0.61 |
| Asian | 92 (2.5%) | 21 (2.6%) | -0.15 | 0.88 |
| Black/African American | 310 (8.6%) | 33 (4.0%) | 4.22 | <0.001 |
| White | 3002 (80.9%) | 707 (86.5%) | -3.78 | <0.001 |
| Other | 300 (8.3%) | 54 (6.6%) | 1.42 | 0.16 |

The PROMIS questionnaires are scaled on a T-scale, with a mean score of 50 and each standard deviation being represented by 10 points. They have been clinically validated in multiple conditions. In particular, the PROMIS questionnaires for fatigue^11-13^, pain interference^14-16^, and social roles^17^ have been validated for syndromes such as fibromyalgia and chronic fatigue syndrome (CFS). These syndromes have similarly diverse, chronic symptoms of incompletely defined etiology similar to the post-COVID syndrome. We chose the PROMIS scales for fatigue, sleep, social roles, physical function and pain based on frequently reported symptoms in post-COVID syndrome both in the literature and in our clinical experience. PROMIS symptom scores >1 standard deviation (SD) of the mean were considered moderate severity and scored >2 SD of the mean were considered severe.

## Results

### Characteristics of respondents

The mean duration between initial positive PCR for SARS-CoV-2 and survey response was 68.4 days. Demographics of the cohort are displayed in Table 1. Notably, the mean age of our respondent population was older than the non-respondents (44 years vs 37 years, p<0.001) and a greater proportion were female (61.1% vs 49.1%, p<0.001). The majority of respondents were primarily English-speaking (96.3%) which was again significantly higher than the proportion in non-respondents (86.3%). Conversely, patients who were not primarily English-speaking made up a lower proportion of respondents compared to non-respondents (interpreter not needed: 1.5% vs 3.7%, p = 0.001; interpreter needed: 2.2% vs 8.1%, p<0.001). The racial composition of the respondent group was also different from the non-respondent group with the proportion of White (86.5% vs 80.9%, p<0.001) and Black patients (4.0% vs 8.6%, p<0.001) reaching statistical significance.

### PROMIS scores

Symptom severity as defined by PROMIS scores is shown in Table 2 and illustrated in Figure 2. There was variation in response rate across the questionnaires ranging from 79.5% completion for the social roles questionnaire to 90.5% on the fatigue questionnaire. PROMIS scores >1 standard deviation worse than normal were noted in social roles (43.2%), pain (17.8%), fatigue (16.2%), physical function (10.6%), and sleep (10.0%). PROMIS scores >2 standard deviations worse than normal were noted in pain (3.8%), fatigue (2.3%), physical function (1.3%), and sleep (1.0%).

**Table 2:** Dysfunction on PROMIS scales in patients recovered from acute COVID-19.

|  | Number of respondents<br>(% of total) | Dysfunction on PROMIS domain (%) |  |  |
| --- | --- | --- | --- | --- |
|  |  | Any (>1 SD) | Moderate (1-2 SD) | Severe (> 2 SD) |
| PROMIS Domain |  |  |  |  |
| Social Roles | 650 (79.6%) | 281 (43.2%) | 281 (43.2%) | 0 (0%) |
| Pain | 738 (90.3%) | 131 (17.8%) | 103 (14.0%) | 28 (3.8%) |
| Fatigue | 739 (90.6%) | 120 (16.2%) | 103 (13.9%) | 17 (2.3%) |
| Physical Function | 719 (88.0%) | 76 (10.6%) | 67 (9.3%) | 9 (1.3%) |
| Sleep | 671 (82.1%) | 67 (10.0%) | 60 (8.9%) | 7 (1.0%) |

**Figure 2:**
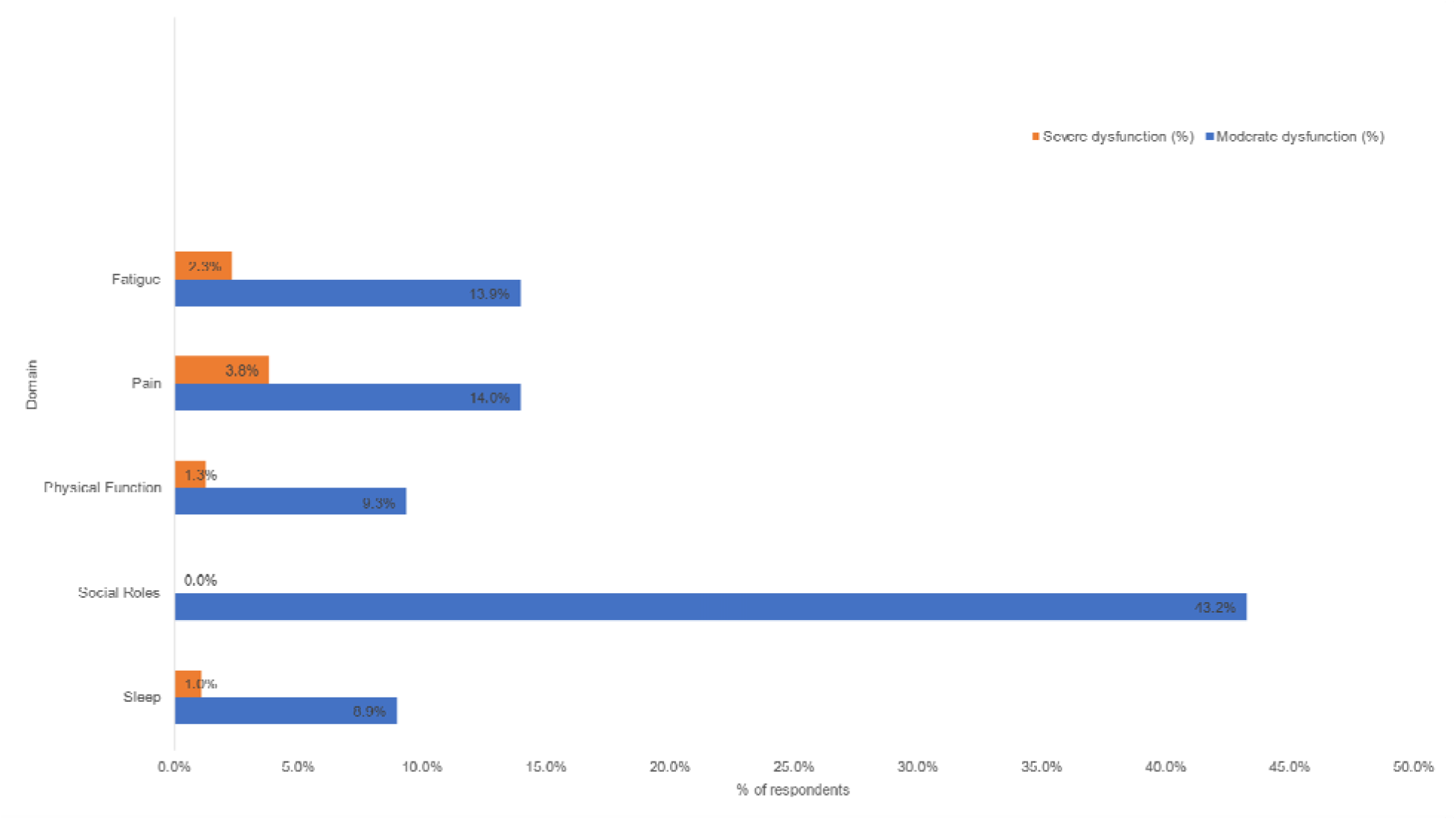
Dysfunction in PROMIS domains in patients recovered from acute COVID-19.

## Discussion

Persistent symptoms (>30 days) were reported by a large proportion of patients after resolution of acute COVID-19 disease. The three most commonly impacted domains of the PROMIS scale were social roles, fatigue, and pain. This is similar to data reported in other studies of post-COVID syndrome which cite pain, fatigue, and dyspnea as the most common manifestations of the condition.^2-4,6,18^These studies did not use the PROMIS scales and instead reported raw symptom data. We believe the current study is the first to utilize the PROMIS scales to assess physical and social dysfunction in patients with post-COVID syndrome, and one of the few to examine the social impact of post-COVID syndrome.

A recent study conducted in the United Kingdom found that fatigue, pain, and social role deficiencies including PTSD symptoms, anxiety/depression, and concentration problems were the among the most common post-discharge symptoms.^3^ Another recent CDC study reported that COVID-19 infections have been associated with adverse mental health conditions including substance use and suicidal ideation.^5^ These findings reflect that patients with post-COVID syndrome are not only more symptomatic, but are impaired in their fulfillment of social roles, which may have further interplay with symptom progression.

One notable difference between our study population and that described in the previous literature is in the age of our demographic. The mean age of our patient population was 44.4 years which is significantly lower than the mean ages previously reported for post-COVID syndrome of 56.5 years^2^ and 70.5 years.^3^ We also report a higher incidence of females in our population than previously reported - 60.5% as compared to 37.1%,^2^ 48.5%,^3^ and 48.1%.^5^ Our average duration of symptoms at the time of survey was 68.4 days, which is significantly longer than the other existing study data for outpatients. The only comparable outpatient study also following outpatients in which a duration of symptoms is available conducted their survey at an average of 16 days following the onset of symptoms.^5^ Additionally, our findings confirm that the post-COVID syndrome may occur across a spectrum of acute illness severity, ranging from those managed outpatient to those severe enough to require hospitalization. We also demonstrate that the duration of post-COVID symptoms in patients who received outpatient management has the potential to extend to a period similar to that of patients who were hospitalized.^2,3^ That the most commonly reported post-discharge symptoms remained congruous suggests a common clinical course of post-COVID syndrome irrespective of age or acute illness severity. We hypothesize that there are intrinsic patient characteristics independent of age and disease severity that may predispose an individual to post-COVID syndrome, and this will need to be elaborated on in future research efforts.

Our sample size is large and heterogeneous, representing both patients who are post-hospitalization as well as those who were never admitted. This compares favorably to previous study sample compositions – 825 patients in our sample compared to 143,^2^ 191,^3^ and 292.^5^ We do however recognize limitations inherent to our cross-sectional study design related to generalizability and inability to assess causation. Another limitation is that there is likely some response bias in that those with more severe symptoms were more likely to respond. The electronic nature of the survey also selects for those with access to the internet and an active portal account which does select against those with low health literacy and socio-economic status. This survey was also delivered in English which may have selected against those with limited English proficiency; however the proportion of participants with limited English proficiency mirrors the population distribution of the Midwest states. The final response rate, however, was 14.7%, which is on the upper end of standard survey response rates for electronic surveys.^19^

## Conclusion

Overall, this brief report presents several new findings regarding the post-COVID syndrome. The PROMIS scales can be reliably used to assess the impact of post-COVID syndrome on patients with findings similar to the published data. The PROMIS social roles score appears to be particularly affected in patients with post-COVID syndrome which is congruent with the impact of this disease on their daily living and interactions. In light of this, we advocate for measuring the social impact of the post-COVID syndrome in future work. It would also be interesting to investigate if PROMIS scales trend towards normal with resolution of symptoms in future projects. We furthermore demonstrate that the post-COVID syndrome has largely preserved symptomatology across a wide variety of ages and illness severity, which may be related to intrinsic patient characteristics that make them more susceptible to the development of post-COVID syndrome. Finally, this report provides a foundation for future exploration of these patient characteristics and potentially providing a standard framework for assessing social and physical dysfunction in affected patients. Better understanding the post-COVID syndrome will help lead to symptom targeted therapies including paced physical rehabilitation techniques and cognitive behavioral therapy.

## Data Availability

Data is maintained by authors and available upon request.

## Notes

Conflict of Interest: None

### Competing Interest Statement

The authors have declared no competing interest.

### Funding Statement

No funding source.

### Author Declarations

Mayo Clinic Institutional Review Board

